# Geospatial Correlation Between COVID-19 Health Misinformation and Poisoning with Household Cleaners in the Greater Boston Area

**DOI:** 10.1101/2020.04.30.20079657

**Authors:** Michael Chary, Daniel Overbeek, Alexandria Papadimoulis, Adina Sheroff, Michele Burns

**Author notes:** Corresponding author: Michael A. Chary, MD PhD, Division of Emergency Medicine, Harvard Medical Toxicology Program, Boston Children’s Hospital, 300 Longwood Ave, Boston, MA 02115.

## Abstract

**Objective:** To determine the relationship between health misinformation on social media and adverse health outcomes.

**Methods:** We analyzed 16,729 calls to the Regional Center for Poison Control and Prevention serving Massachusetts and Rhode Island (MARI PCC) and 25,231 tweets discussing treating COVID-19 with house cleaners.

**Results:** Half of the spikes in calls to MARI PCC about exposures to cleaners were preceded 2–3 days earlier by tweets advocating ingesting or insufflating bleach to cure COVID-19. The relationship was only statistically significant for tweets in the Greater Boston Area and calls to MARI PCC. [Results of network analysis].

**Conclusions:** Health misinformation on social media had a spatiotemporally specific relationship with increased calls to Poison Control and referrals to hospitals for cleaner ingestions. The spatiotemporal specificity of our results strengthens our conviction that the increased calls were driven in part by health misinformation on social media.

**Public Health Implications:** Health misinformation can directly lead to secondary harm. Thematically targeted messages at specific loci in the social network may stem this harm.

## Introduction

Prolific social media activity has accompanied the COVID-19 pandemic^1^. Health misinformation on social media may lead to harm from using toxic substances^2^, especially in a pandemic where few vetted therapies exist. Some remedies proposed in the media (e.g., gargling bleach, mixing cleaners) are toxic even at minimal doses. Analyses of social media have tracked the spread of infectious diseases^3^, but have not investigated harm from unapproved treatments for an infectious disease. We found a relationship between social media discussions about treating COVID-19 and adverse health outcomes. We hypothesized and observed that discussions of untested remedies on social media would lead to the public to try those remedies and then experience adverse reactions and toxicity. As a result, an increase in poisonings reported to Poison Control Centers (PCCs) would increase.

## Methods

This study was granted exemption by the Institutional Review Board because it involved only analyses of publicly available data or secondary analyses of deidentified data already gathered.

### Data Acquisition

We obtained call logs from the Regional Center for Poison Control and Prevention Serving Massachusetts and Rhode Island (MARI PCC) between December 1, 2019 and March 15, 2020, and for the same time period for four prior years. We included all calls involving household cleaning products. This category, as defined by the American Academy of Clinical Toxicologists, includes ammonia-, bleach-, and borate-based cleaners as well as phenol, and pine oil. We acquired publicly available tweets that (1) were emitted from the BMA and (2) contained keywords related to coronavirus (i.e., covid-, coronavi-), cleaning products (i.e., bleach, ammonia, hypochlorite, borate), and health claims (cure, treat). We considered tweets as emitted from Greater Boston Area if they fell within the boundaries establish by the Metropolitan Area Planning Council.

### Time Series Analysis

We downsampled tweets to daily frequency. We then detrended each time series, daily PCC call volume and tweet counts, by taking the first difference of the logarithms of the time series and then scaled each time series to be between 0 and 1. The first difference of a time series (X_1_, X_2_, X_3_, X_4_, …) is the series (X_2_-X_1_, X_3_-X_2_, X_4_-X_3_, …). Taking the first difference is one way to account for nonstationarity. After this, both time series were stationary (augmented Dickey-Fuller tests less than critical values). We calculated the correlation between the differenced scaled time series.

### Results

MARI PCC received 16,729 calls between December 1, 2019-March 15, 2020 (“COVID-19 period”), an increase of 9.9% from 14,897 calls during the same time period the previous year, and 5.6% over the 5-year median. Calls about cleaners during the COVID period exceeded the 95% confidence interval for the 5 year-median. Calls about hand sanitizer, acetaminophen, and total calls did not change. 143/1047 (13.5%) calls regarding cleaners required referral to the hospital compared to 146/1092 (16.7%) in December 1, 2018-March 15, 2019. We obtained 25,231 unique tweets from BMA discussing cleaning products. Of those tweets, 13,362 mentioned coronavirus and cleaner keywords; 2,416 mentioned all 3 classes of keywords. The pattern of daily tweets mentioning all 3 classes of keywords anticipated 30% of the dynamics of calls 2–4 days later to MARI PCC (Figure 1). The peaks in February and March 2020 correspond to reactions to QAnon, a far-right conspiracy group, stating that gargling bleach could cure coronavirus^4^. This correlation was significant for tweets from BMA containing keywords from all categories.

**Figure 1.**
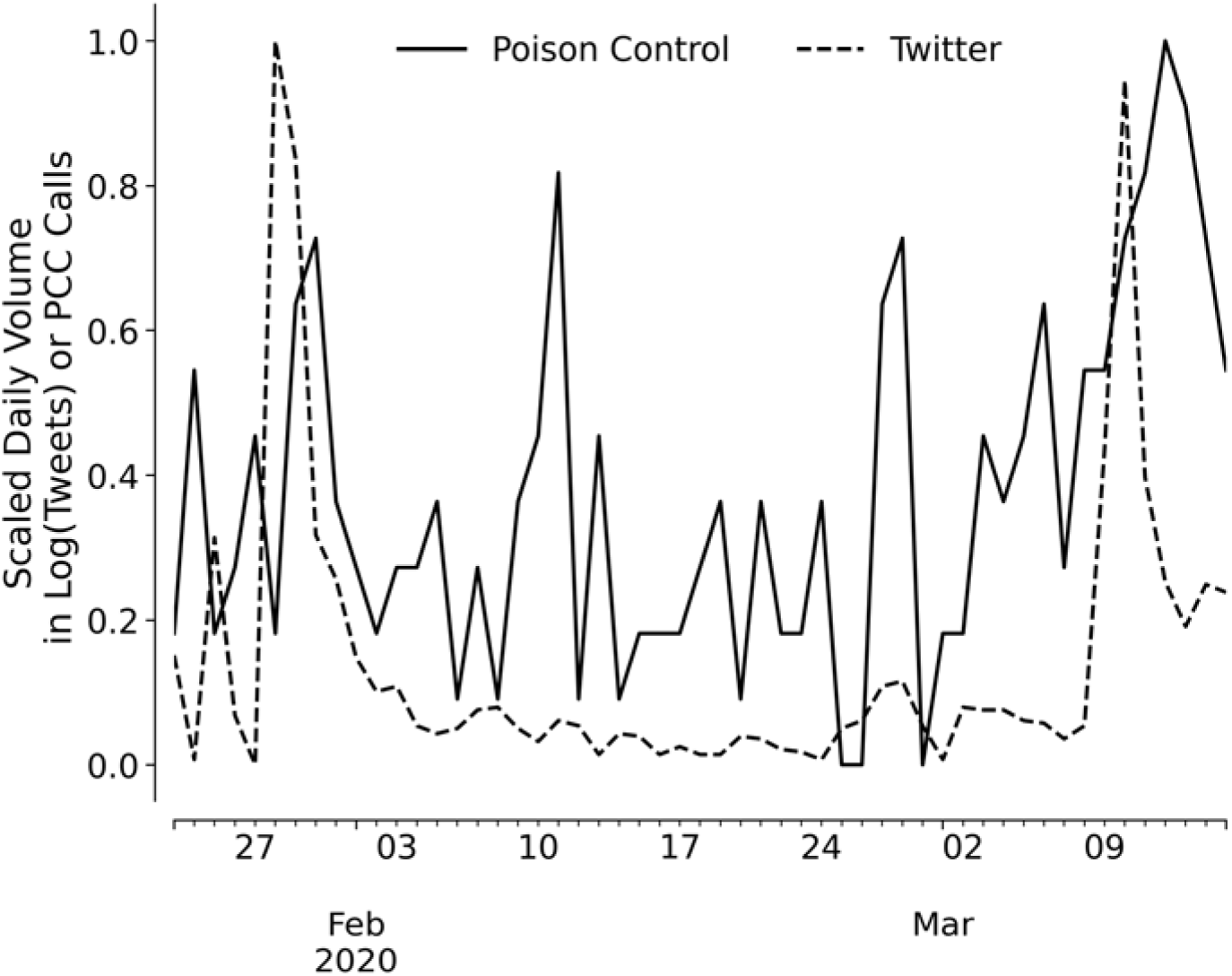
Time course and cross-correlation function between tweets about cleaners and calls to PCC about cleaning products. **Left panel:** X-axis displays the date by days. Y-axis shows ddaily count of calls to MARI PCC (solid line) and logarithm of tweets (dashed line).

### Public Health Relevance

Online discussions about untested popularized treatments for COVID-19 can predict the dynamics of calls to PCCs 3–4 days later. Our findings suggest that online discussions directly inform behavior that leads to adverse health outcomes. 20% of Americans use Twitter as a platform for information and communication^5^. Our results suggest that health misinformation on social media can have a temporal relationship with secondary harm. We identified this relationship between social media and poisonings prior to the misinformation the week of April 19 on injecting bleach. We are concerned that this discussion will promote secondary harm as with the two prior episode. MARI PCC is in Boston, Massachusetts; 4 out of 5 people in Massachusetts live in the Boston Metropolitan Area (BMA).

## Data Availability

All supporting code is available upon request. Deidentified versions of underlying data may be available on an individual basis.

## Supplmental

**Supplmental Table 1.** Call Volume to MARI PCC from Dec 2019 - Mar 2020 and same time period from four years prior. IQR, Interquartile Range.

| Substance | Median No. of Calls (± IQR)<br>for Dec-Mar over Last 5 Years | No. of Calls Dec 2019-Mar<br>2020 | Change in calls (%) |
| --- | --- | --- | --- |
| All Calls | 15,516 ± 447 (14,812-16,219) | 16,729 | 7.3% |
| Cleaners | 258 ± 12 (225-291) | 305 | 15.4% |
| Hand Sanitizers | 1,047 ± 36 (968-1,126) | 1,092 | 4.1% |
| Acetaminophen | 579 ± 25 (531-627) | 608 | 4.8% |

## Notes

### Competing Interest Statement

The authors have declared no competing interest.

### Funding Statement

No specific funding was received for this project.

